# Effectiveness of Four Vaccines in Preventing SARS-CoV-2 Infection in Kazakhstan

**DOI:** 10.1101/2022.04.14.22273868

**Authors:** Dilyara Nabirova, Roberta Horth, Manar Smagul, Gaukhar Nukenova, Aizhan Yesmagambetova, Daniel Singer, Alden Henderson, Alexey Tsoy

## Abstract

**BACKGROUND:** In February 2021 Kazakhstan began offering COVID-19 vaccines to adults. Breakthrough SARS-CoV-2 infections raised concerns about real-world vaccine effectiveness. We aimed to evaluate effectiveness of four vaccines against SARS-CoV-2 infection.

**METHODS:** We conducted a retrospective cohort analysis among adults in Almaty using aggregated vaccination data and individual-level breakthrough COVID-19 cases (≥14 days from 2^nd^ dose) using national surveillance data. We ran time-adjusted Cox-proportional-hazards model with sensitivity analysis accounting for varying entry into vaccinated cohort to assess vaccine effectiveness for each vaccine (measured as 1-adjusted hazard ratios) using the unvaccinated population as reference (N=565,390). We separately calculated daily cumulative hazards for COVID-19 breakthrough among vaccinated persons by age and vaccine month.

**RESULTS:** From February 22 to Sept 1, 2021 in Almaty, 747,558 (57%) adults were fully vaccinated (received 2 doses) and 108,324 COVID-19 cases (11,472 breakthrough) were registered. Vaccine effectiveness against infection was 78% (sensitivity estimates: 74–82%) for QazVac, 77% (72– 81%) for Sputnik V, 71% (69–72%) for Hayat-Vax, and 69% (64–72%) for CoronaVac. Among vaccinated persons, the 90-day follow-up cumulative hazard for breakthrough infection was 2.2%. Cumulative hazard was 2.9% among people aged ≥60 years versus 1.9% among persons aged 18–39 years (p<0.001), and 1.2% for people vaccinated in February–May versus 3.3% in June–August (p<0.001).

**CONCLUSION:** Our analysis demonstrates high effectiveness of COVID-19 vaccines against infection in Almaty similar to other observational studies. Higher cumulative hazard of breakthrough among people >60 years of age and during variant surges warrants targeted booster vaccination campaigns.

**What is already known on this topic:**

- Plenty of data are published on effectiveness of mRNA vaccines; however, these vaccines were not widely available in many low- and middle-income countries in 2021.
- There are no real-world effectiveness studies on several vaccines available in the Central Asia region, including QazVac vaccine, an inactivated vaccine developed by Kazakhstan.
- Understanding how these vaccines are performing outside of clinical trials is critical for the COVID-19 response and lack of published data can contribute to vaccine hesitancy.

**What this study adds:**

- Our study demonstrated that at the population-level the four vaccines against COVID-19 used in Kazakhstan were effective at preventing SARS-CoV-2 infection.
- Vaccination reduced the risk of infection by 76% and prevented over 100,000 cases of SARS-CoV-2 infection in Almaty, the country’s most populous city.
- This is also the first study that demonstrated high vaccine effectiveness in real-world conditions of QazVac, developed in Kazakhstan.

**How this study might affect research, practice or policy:**

- Policy makers in Kazakhstan and the Central Asia region need data on vaccines provided in the region to update evidence-based vaccine guidelines for different populations.

## INTRODUCTION

From March 2020 to December 17, 2021 in Kazakhstan, over one million people were diagnosed with coronavirus disease 2019 (COVID-19); ∼18,000 died.^1^ Vaccines have become a key tool for reducing COVID-19-associated morbidity and mortality and preventing transmission of SARS-CoV-2. In February 2021, Kazakhstan began offering COVID-19 vaccines to adults aged 18 years and older. By December 9, 2021, 71% of adults (∼eight million people) had received at least two doses of vaccines, including Sputnik V (Gam-COVID-Vac, Gamaleya Research Institute of Epidemiology and Microbiology, Russia), QazVac (QazCovid-in, Research Institute for Biological Safety Problems, Kazakhstan), CoronaVac (Sinovac Biotech, China), Sinopharm (BBIBP-CorV, Beijing Institute of Biological Products, China) and Hayat-Vax (BBIBP-CorV, G42 Healthcare, United Arab Emirates).^2^

Of the five vaccines available in Kazakhstan, two (CoronaVac and Sinopharm) were listed for emergency use globally by the World Health Organization (WHO) by end of 2021. The WHO pre-qualifies and lists vaccines based on several guidelines^3^ including Good Manufacturing Practice (GMP) and >50% vaccine efficacy in preventing SARS-CoV-2 outcomes in clinical trials.^4, 5^ Among the WHO listed vaccines, vaccine efficacy against symptomatic disease in randomized-controlled trials was 51-84% for CoronaVac, 79% for Sinopharm, 63-76% for AstraZeneca, and >90% for Moderna and Pfizer–BioNTech vaccines.^4, 5,6^

Although not listed by the WHO, Sputnik V and QazVac vaccines have received emergency use authorization from the Kazakhstan Ministry of Health. They have shown interim efficacy of >90% and safety in phase III and II clinical trials, respectively.^7-10^ Additionally, Sputnik V demonstrated 79-86% effectiveness against infection and 85-93% effectiveness against COVID-19 related death in a number of observational studies outside of clinical trials (real-world).^11, 12^

Increasing breakthrough COVID-19 infections among vaccinated people raised concerns about the real-world effectiveness of available vaccines. By the end of 2021, there were no real-world effectiveness studies on the QazVac vaccine and limited data on the effectiveness of Sputnik V. Understanding how vaccines are performing outside of clinical trials is important for healthcare system preparedness and mitigation strategies. Policy makers in Kazakhstan needed data to create evidence-based vaccine guidelines, including policies for booster vaccine doses for different populations. We aimed to estimate daily hazard of breakthrough SARS-CoV-2 infection among vaccinated people in Almaty, to identify groups at higher risk for breakthrough infection, and to evaluate the overall effectiveness of four vaccines against infection.

## METHODS

### Study design

We conducted a retrospective, observational population-based cohort study of the vaccine effectiveness of four different vaccines against SARS-CoV-2 infection among adults 18 years or older in Almaty, Kazakhstan. Almaty is the economic capital and largest city of Kazakhstan with an adult population of 1,313,040 (∼10% of the national adult population).

### Key definitions

A breakthrough COVID-19 infection was defined as having confirmed or probable COVID-19, including people with asymptomatic disease, ≥14 days after receiving the 2^nd^ dose of a COVID-19 vaccine.

A confirmed COVID-19 case was defined as the detection of SARS-CoV-2 ribonucleic acid (RNA) in a respiratory specimen collected from a person aged ≥18 years, including those with asymptomatic disease.

A probable COVID-19 case was a patient who met clinical criteria (chest imaging suggesting COVID-19, anosmia, ageusia in the absence of any other identified cause), did not have a positive SARS-CoV-2 PCR test, and was a contact of a probable or confirmed case or linked to a COVID-19 cluster.

Fully vaccinated was defined as having received two COVID-19 vaccine doses (there were no single dose COVID-19 vaccines offered in Kazakhstan). Unvaccinated was defined as never having received any COVID-19 vaccine. Partial vaccination was having received only one vaccine dose. People who received only one dose were excluded from analysis.

We classified people into four groups. People who were:

1. unvaccinated (never received any COVID-19 vaccine) and diagnosed with COVID-19,
2. unvaccinated (never received any COVID-19 vaccine) and not diagnosed with COVID-19,
3. fully vaccinated (two doses) and diagnosed with breakthrough COVID-19, and
4. fully vaccinated (two doses) and not diagnosed with breakthrough COVID-19.

### Study population

Our analysis included Almaty residents ages 18 years and older. The following people were excluded from analysis (Supplementary Figure 1):

- People that received only their first dose of vaccine and had a positive SARS-CoV-2 PCR test <14 days after the first vaccine dose.^13^
- People whose records were inconsistent with national vaccination guidelines (e.g. second dose was <7 days of the first dose or they had COVID-19 diagnosis <25 days before their second dose).
- People who received the Sinopharm vaccine (only 20 people).
- People who had SARS-CoV-2 PCR positive result <14 days after their second dose. This last group was included in sensitivity analysis.

Data on history of COVID-19 prior to the study period was not available; therefore, we do not exclude based on prior disease.

### Data sources

Individual-level data on COVID-19 cases among anyone who had ever received a COVID-19 vaccine were provided by the Emergency Operational Center (EOC) of the National Center of Public Health of the Ministry of Health in Kazakhstan from February 1 to September 1, 2021. Data on all COVID-19 cases from testing centers, laboratories, hospitals, and healthcare providers are collected by the EOC. This clinical record dataset included age, comorbidities, dates of COVID-19 vaccination, vaccine name, date of SARS-CoV-2 PCR test result or diagnosis, hospitalization, death, and disease severity. Disease severity was based on the WHO Scale adapted by the Kazakhstan clinical protocol of diagnosis and treatment. People with critical disease had acute respiratory distress, those with severe disease had severe pneumonia, those with moderate disease had pneumonia, and those with mild disease did not have pneumonia. Data aggregated per day on SARS-CoV-2 polymerase chain reaction (PCR) testing, which included the number of people testing positive per day for the same period, were also provided.

Aggregated data on vaccination were received from the Salidat Kairbekova National Research Center for Health Development of the Ministry of Health in Kazakhstan. Variables were aggregated by date and included the number of people vaccinated per day per age group and vaccine type.

### Statistical analysis

Analyses were performed using R statistical software, version 4.0.3 (R Foundation for Statistical Computing). We calculated and plotted attack rates for the vaccinated and unvaccinated populations for each day of our analysis for each vaccine type. We fitted time-dependent Cox proportional hazards regression models using the coxph() function of the survival package to assess the time to COVID-19 diagnosis by vaccination status. Vaccine status was a time-dependent covariate that changed when the person received their 1st dose and 14 days after their 2^nd^ dose.

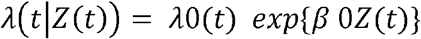

The hazard at time t depends on the value of covariates at time Z(t). The regression effect of Z(·) is constant β over time. The adjusted hazard ratio was exp{β 0Z(t)}. Adjusted vaccine effectiveness, measured as 1-exp{β 0Z(t)}, and adjusted for day and vaccine status, was assessed for each of the 4 vaccines using the unvaccinated population as the reference group.

The time of entry into the model was set at February 1, 2021, when COVID-19 vaccines became available in Almaty. The entire population of adults ≥18 years old in Almaty (N=1,313,040) was classified into the unvaccinated group at-risk for COVID-19 group. People contributed person-time to the unvaccinated population until they either received a COVID-19 diagnosis or a COVID-19 vaccine. After people received their first vaccine, they did not contribute person-time to either population (vaccinated or unvaccinated) from their first dose to 14 days after their second dose after which they contributed person-time to the vaccinated population. We performed sensitivity analysis by changing the dates at which people entered the vaccinated population (at first dose, second dose, second dose + 7 days or second dose + 14 days) and the dates of definition of a COVID-19 breakthrough case (after second dose, second dose + 7 days or second dose + 14 days). We show sensitivity estimates throughout for our results to more accurately express the uncertainty around estimates.

We separately calculated crude vaccine effectiveness for each vaccine using the following formula:

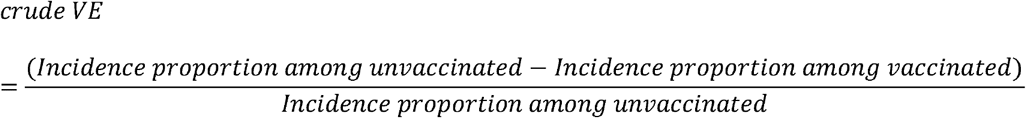

We also estimated the number of COVID-19 cases prevented by multiplying the number of vaccinated people by the overall incidence among the unvaccinated population, and subtracting the number of breakthrough cases.

Lastly, we conducted sub analysis on the vaccinated population, we calculated the cumulative hazard percent of getting COVID-19 using survfit() and plotted Kaplan-Meyer cumulative hazard curves using ggsurvplot() function and by vaccine name, age group, and month. The log-rank test was used to test difference in hazard curves.

## RESULTS

From February 22 to September 1, 2021 in Almaty, 747,558 residents aged ≥18 years were fully vaccinated against COVID-19 with Sputnik-V (668,452), Hayat-Vax (42,437), CoronaVac (22,181), or QazVac (14,488) and met inclusion criteria (supplement figure). During this time, Almaty had 108,324 probable and confirmed COVID-19 cases among people ≥18 years old, including 23,858 cases among people with at least one COVID-19 vaccination record (supplement figure). Of these 11,472 were fully vaccinated and met criteria for breakthrough SARS-CoV-2 infection.

Among fully vaccinated people with breakthrough SARS-CoV-2 infections (table 1), 24% were ≥ 60 years old and 11% had a known comorbidity and presence of comorbidity was unknown for 72%. The median interval between vaccine doses was 21 days (range 8–166). The median interval between second dose and breakthrough infection was 59 days (range 14–178). Disease was classified as critically severe for 0.7% and severe for 6.8% of cases, and 29% were hospitalized. There were 120 deaths (1%).

**Table 1.** Characteristics of fully vaccinated people with SARS-CoV-2 breakthrough infection, Almaty, Kazakhstan, 2021

| Characteristics | N=11,472 | % or (range) |
| --- | --- | --- |
| Sex |  |  |
| Female | 6,923 | 60.3 |
| Male | 4,549 | 39.7 |
| Age, median (range)<sup>o</sup> | 45 | (range 18 to 96) |
| 18-39 years | 4,360 | 38.0 |
| 40-59 years | 4,351 | 37.9 |
| ≥ 60 years | 2,761 | 24.1 |
| Had a comorbidity |  |  |
| No | 1,892 | 16.5 |
| Yes | 1,271 | 11.1 |
| Unknown | 8,309 | 72.4 |
| Fully vaccinated |  |  |
| Sputnik V | 9,826 | 85.7 |
| Hayat-Vax | 955 | 8.3 |
| QazVac | 187 | 1.6 |
| CoronaVac | 504 | 4.4 |
| COVID-19 waves in Kazakhstan |  |  |
| February 1- June 30, 2021 | 402 | 3.5 |
| July 1- 31, 2021 | 2,335 | 20.4 |
| August 1- September 1, 2021 | 8,735 | 76.1 |
| PCR-identified cases |  |  |
| PCR - | 560 | 4.9 |
| PCR + | 10,912 | 95.1 |
| Interval between 1st and 2nd dose in days, median (range) | 21 | (range 8 to 166) |
| 8-14 days | 259 | 2.3 |
| 15-19 days | 395 | 3.4 |
| 20-25 days | 8,449 | 73.6 |
| 26-46 days | 1,906 | 16.6 |
| 47-166 days | 463 | 4.0 |
| Days since fully vaccinated to disease onset | 59 | (range 14 to 178) |
| COVID-19 severity* |  |  |
| Critical disease | 78 | 0.7 |
| Severe disease | 776 | 6.8 |
| Moderate disease | 2,154 | 18.8 |
| Mild disease | 8,464 | 73.8 |
| Hospitalized |  |  |
| No | 8,146 | 71.0 |
| Yes | 3,326 | 29.0 |
| Number of hospitalizations** |  |  |
| One hospitalization | 3,285 | 98.8 |
| Two hospitalizations | 41 | 1.2 |
| Length of hospitalization in days, median (range)** | 9 | (range 1 to 35) |
| Outcome |  |  |
| Recovered or transferred | 11,352 | 99.0 |
| Died | 120 | 1.0 |

\* WHO Scale adapted by the Kazakhstan clinical protocol of diagnosis and treatment (Coronavirus infection COVID-19 in adults);\*\*Subset of persons having been hospitalized

Vaccination began on February 1, 2021 for Sputnik, April 25 for QazVac, May 3 for Hayat-Vax and June 6 for CoronaVac. During the study period, the adjusted vaccine effectiveness for all four vaccines was 76% (low and high sensitivity percent range of 71.1–80.0%) (table 2). Adjusted vaccine effectiveness varied by vaccine and was 77.0% (71.4–80.8%) for Sputnik V, 78.6% (74.2–82.0%) for QazVac, 71.2% (69.3–73.0%) for Hayat-Vax, and 69.5% (64.7–71.5%) for CoronaVac (p<0.001).

**Table 2.** Vaccine effectiveness against SARS-CoV-2 infection of four COVID-19 vaccines in Almaty, Kazakhstan, 2021

| Category | N | % aVE | 95% CI | Sensitivity analysis |  |  |  | % cVE | 95% CI |
| --- | --- | --- | --- | --- | --- | --- | --- | --- | --- |
|  |  |  |  | Low | 95% CI | High | 95% CI |  |  |
| Vaccine status |  |  |  |  |  |  |  |  |  |
| Vaccinated | 747,558 | 76.4* | (75.9-76.9) | 71.1 | (70.5-71.6) | 80.0 | (79.6-80.4) | 89.7 | (89.5-89.9) |
| Not vaccinated | 565,370 | Ref |  |  |  |  |  | Ref |  |
| Vaccine name |  |  |  |  |  |  |  |  |  |
| QazVac | 14,488 | 78.6* | (75.3-81.4) | 74.2 | (70.2-77.7) | 82.0 | (79.2-84.4) | 85.8 | (83.6-87.7) |
| Sputnik-V | 668,452 | 77.0* | (76.5-77.5) | 71.4 | (70.8-72.1) | 80.8 | (80.4-81.2) | 90.4 | (90.2-90.6) |
| Hayat-Vax | 42,437 | 71.2* | (69.3-73.0) | 69.1 | (67.1-71.0) | 72.4 | (70.6-74.1) | 73.2 | (71.4-74.8) |
| CoronaVac | 22,181 | 69.5* | (66.7-72.0) | 64.7 | (61.5-67.7) | 71.5 | (68.9-73.9) | 71.1 | (68.5-73.5) |
| Not vaccinated | 565,370 | Ref |  |  |  |  |  | Ref |  |

\*Time-dependent Cox-proportional hazards log-rank test P-value <0.001
aVE: adjusted vaccine effectiveness, adjusted for time and vaccine status
CI: Confidence Interval
Low: day of contribution of person-time to vaccinated population begins on day of 1st dose
High: day of contribution of person-time to vaccinated population begins 14 days after 2nd dose
cVE: crude vaccine effectiveness; persons partially vaccinated (n=7482) and those with a case registered
before February 22, 2021 (n=2018) were excluded from the unvaccinated reference group

For all vaccines, the attack rate for COVID-19 per 100,000 was consistently higher over the 7-day moving average for unvaccinated persons than for vaccinated persons (figure 1). The 7-day moving average of daily vaccine effectiveness for all four vaccines remained above the WHO 50% threshold for vaccine efficacy^5^, except for one-week dips for QazVac in June and for CoronaVac and Hayat-Vax in August. Based on overall incidence in the study period, vaccination with these four vaccines prevented 100,213 cases of SARS-CoV-2 breakthrough infection in Almaty.

**Figure 1.**
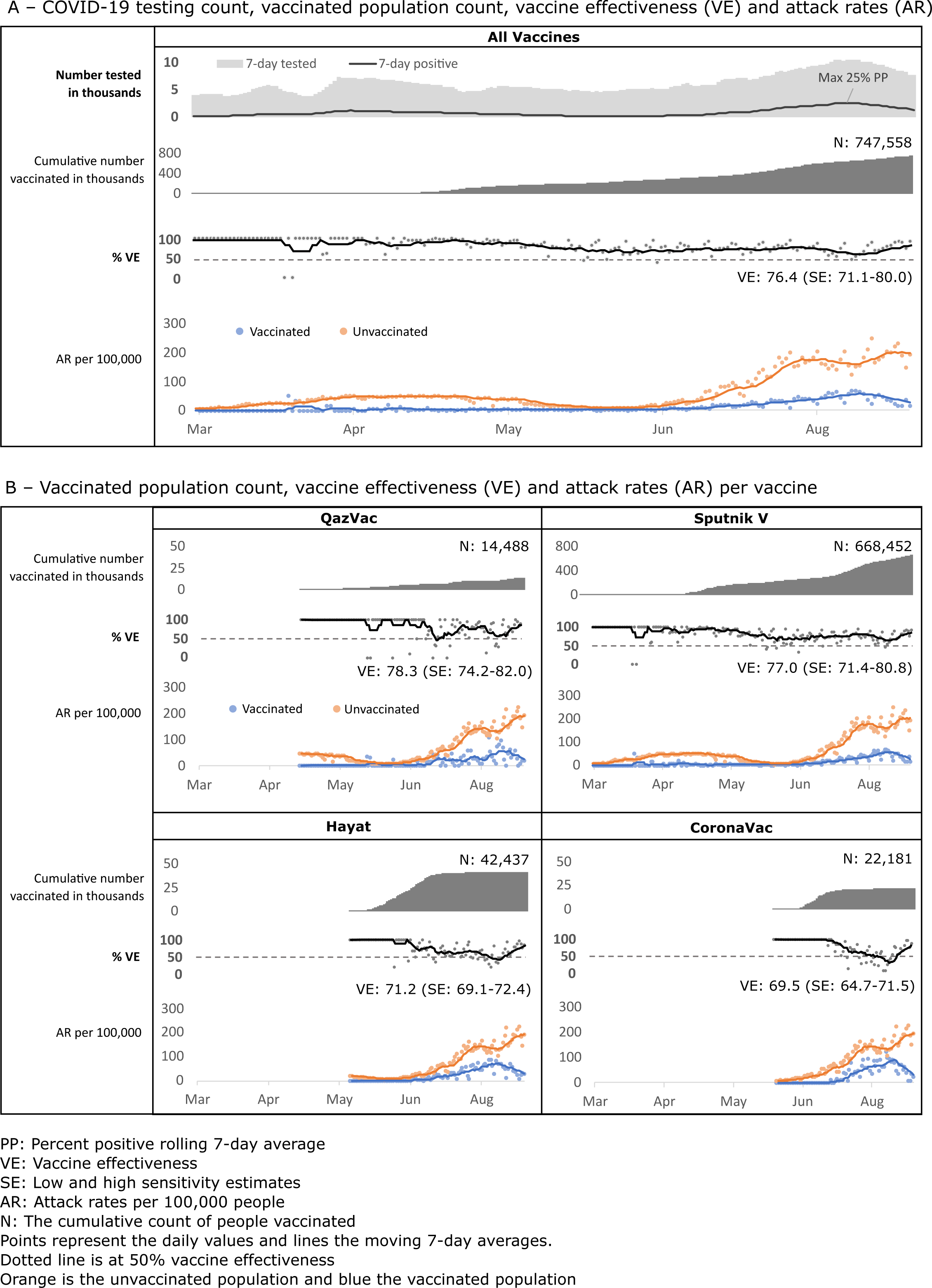
Cumulative vaccinated, vaccine effectiveness against SARS-CoV-2 infection, and attack rates among vaccinated and unvaccinated populations by four COVID-19 vaccines in Almaty, Kazakhstan, 2021

Among people who were fully vaccinated (n=747,558), the 90-day cumulative hazard of developing a SARS-CoV-2 breakthrough infection was 2.2%, ranging from 2.0% for Sputnik V to 3.5% for Hayat-Vax (table 3). The cumulative hazard was significantly different by age group. For people 60 years old and older the cumulative hazard was double that of people 18–39 years old at the 180-day (12.8% vs 5.0%, respectively) and 90-day follow-ups (2.9% vs 1.9%).

**Table 3.** Cumulative hazard of SARS-CoV-2 breakthrough infection among persons fully vaccinated by four COVID-19 vaccines, Almaty, Kazakhstan, 2021

| Group | N | Cumulative Hazard Percent |  |  |  |  |  |  | Trend Line | P-value |
| --- | --- | --- | --- | --- | --- | --- | --- | --- | --- | --- |
|  |  | Day | 30 | 60 | 90 | 120 | 150 | 180 |  |  |
| All vaccine | 747 558 |  | 0.5% | 1.2% | 2.2% | 3.9% | 5.1% | 6.7% |  | <0.001 |
| Sputnik | 668 452 |  | 0.4% | 1.0% | 2.0% | 3.7% | 4.9% | 6.5% |  |  |
| QazVac | 14 488 |  | 0.5% | 1.4% | 2.5% | 3.5% |  |  |  |  |
| Hayat | 42 437 |  | 0.4% | 2.0% | 3.5% |  |  |  |  |  |
| CoronaVac | 22 181 |  | 1.0% | 2.6% |  |  |  |  |  |  |
| Age |  |  |  |  |  |  |  |  |  | <0.001 |
| 18-39 | 387 024 |  | 0.4% | 1.1% | 1.9% | 2.9% | 3.8% | 5.0% |  |  |
| 40-59 | 272 225 |  | 0.5% | 1.2% | 2.2% | 3.9% | 5.2% | 6.8% |  |  |
| 60+ | 88 309 |  | 0.5% | 1.4% | 2.9% | 5.9% | 8.8% | 12.8% |  |  |
| Month |  |  |  |  |  |  |  |  |  | <0.001 |
| Feb | 1 171 |  | 0.1% | 0.3% | 0.8% | 0.9% | 1.3% | 2.7% |  |  |
| Mar | 2 844 |  | 0.3% | 0.5% | 0.6% | 1.0% | 2.1% | 3.8% |  |  |
| Apr | 53 371 |  | 0.1% | 0.2% | 0.7% | 2.3% | 3.6% |  |  |  |
| May | 147 046 |  | 0.1% | 0.3% | 1.3% | 3.0% |  |  |  |  |
| Jun | 105 171 |  | 0.3% | 1.5% | 2.7% |  |  |  |  |  |
| Jul | 221 353 |  | 0.8% | 2.0% |  |  |  |  |  |  |
| Aug | 208 821 |  | 0.6% |  |  |  |  |  |  |  |
| Sep | 7 781 |  |  |  |  |  |  |  |  |  |

This divergence in hazard by age group over follow-up time was observed across the four vaccines (p<0.001) (figure 2). Cumulative hazard curves began to diverge by age at approximately 90 days postvaccination for Sputnik V, and at approximately 30 days postvaccination for other vaccines. Cumulative hazard is also significantly different by study period (p<0.001) when comparing people who were vaccinated in March to May 2021 to those vaccinated in June to September 2021 (the period when the SARS-CoV-2 Delta variant was active in the region). However, in vaccine-specific analysis, this difference was only significant for QazVac and Sputnik V.

**Figure 2.**
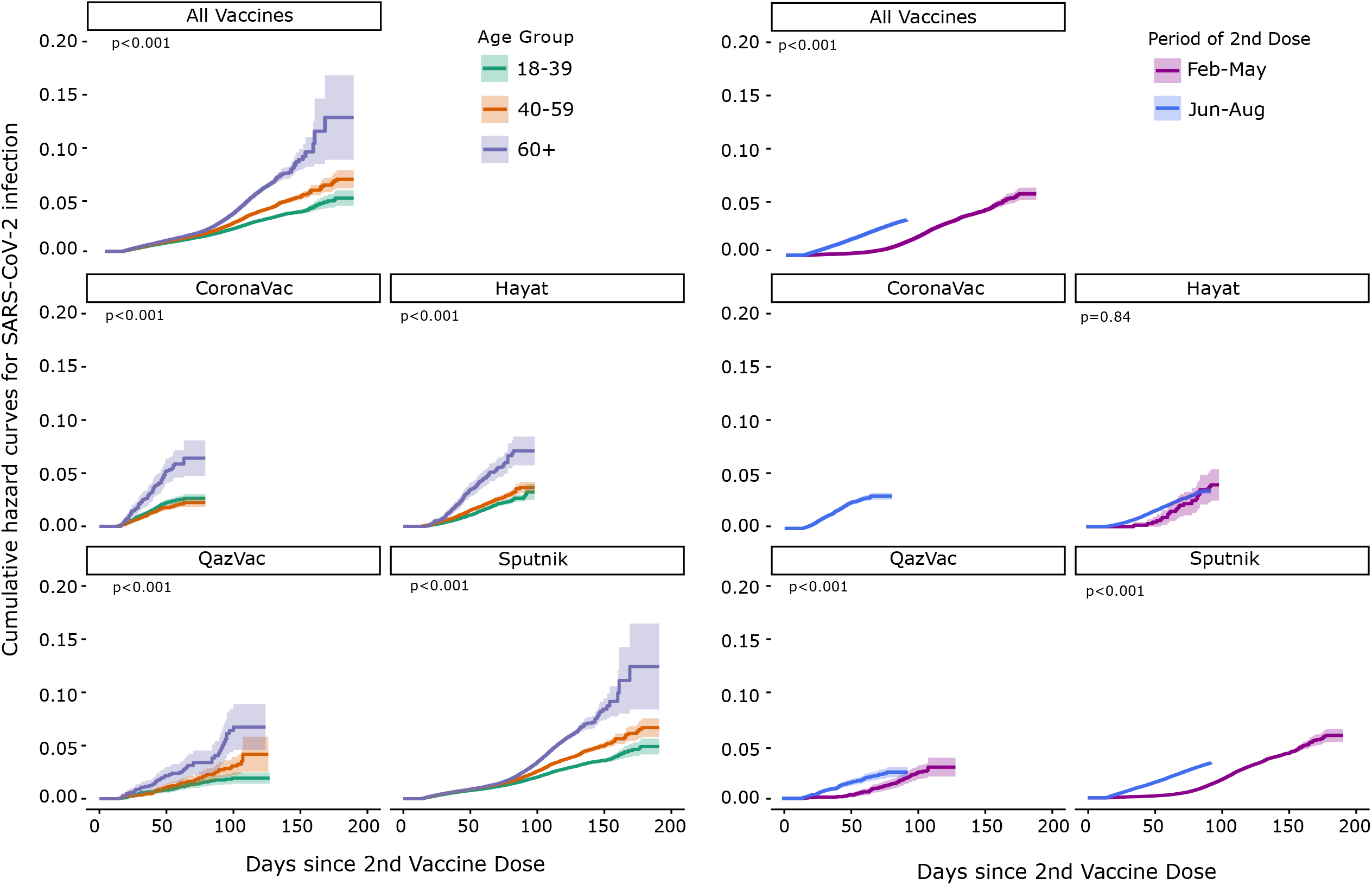
Cumulative hazard curves by age group and period of 2^nd^ dose by vaccine type by 4 COVID-19 vaccines, Almaty, Kazakhstan, 2021

## DISCUSSION

Our study demonstrated that at the population-level the four vaccines against COVID-19 used in Kazakhstan were effective at preventing SARS-CoV-2 infection. Vaccination reduced the risk of infection by 76% and prevented over 100,000 cases of SARS-CoV-2 infection in Almaty, the country’s most populous city. The vaccine effectiveness for all four vaccines exceeded the 50% vaccine efficacy minimum threshold set by the WHO for approving vaccines. Vaccine effectiveness also remained relatively stable over the period, despite a slight decrease observed during the months when the SARS-CoV-2 Delta variant became the predominant circulating strain worldwide.

QazVac had 79% vaccine effectiveness in our study, but accounted for only 2% of people vaccinated. Vaccine effectiveness for QazVac in our analysis was similar to the vaccine efficacy reported in phase III clinical trials (81%).^10^

The predominant vaccine given in Kazakhstan during the period of analysis was Sputnik V (89% of vaccinated people) and our observed vaccine effectiveness of 73% compares to (79% and 86%) vaccine effectiveness from limited observational studies, but lower than 92% vaccine efficacy estimate from phase III clinical trials. However, these clinical trials took place before Delta became the predominant variant.^8, 11, 12^

CoronaVac had the lowest vaccine effectiveness in our analysis, and the estimate fell within the range of vaccine efficacy phase III trials (51% to 84%) in several countries.^6, 14-17^ It is also consistent with vaccine effectiveness against symptomatic disease reported in a comparable study from Chile (66%).^18^ Vaccine effectiveness for Hayat-Vax (71%) was also within the range of vaccine effectiveness reported in similar studies in Hungary and Peru (69% and 50%, respectively)^11, 19^ and vaccine efficacy reported by Sinopharm (the same BBIBP-CorV vaccine manufactured in China) for phase III clinical trials (79%).^6, 20^

Although COVID-19 vaccines used globally are highly effective, people who are fully vaccinated may experience breakthrough SARS-CoV-2 infections, especially when the incidence is high, and the proportion of people vaccinated in a population is low. In Kazakhstan, population vaccination began shortly before the Delta surge. As expected, breakthrough cases occurred among vaccinated people, increasing as SARS-CoV-2 incidence increased in the country. However, out of every 100 people fully vaccinated in Almaty during our study period less than 2 had breakthrough infections. The low proportion of people getting breakthrough COVID-19 in our study is within the range of that reported in other settings (<1 to 8%).^21, 22^ Not only are vaccines effective for reducing infection, they are also highly effective at reducing morbidity and mortality; ^23, 24^ in our study, the majority of people with breakthrough infection had mild disease.

Proportion of breakthrough infections increased over time, especially among people >60 years of age and during the SARS-CoV-2 Delta variant surge.^6, 25^ This increase was most notable among people vaccinated with Sputnik V because it had the longest period of follow-up, but the follow-up period was limited to <90 days for 2 of the 4 vaccines studied. We cannot distinguish whether the declining effectiveness was attributable to waning immunity or emergence of new SARS-CoV-2 variants. However, waning immunity has been demonstrated in several studies that found reduced neutralizing antibodies over time in people vaccinated against COVID-19 with CoronaVac, Sputnik V, and BNT162b2 (Hayat-Vax) vaccines, and others.^6, 26^ Our results align with Kazakhstan’s recommendation to offer booster vaccinations for older populations, and with CDC and WHO recommendations for booster doses for all vaccinated adults.

During data cleaning, we found 1,204 cases where the dates of vaccination and disease onset conflicted with national vaccine administration directives. While some errors could be recording problems, many are likely falsified certification of vaccinations (fake vaccine passports). Fake vaccine passports are known problems in the Central Asia Region and in Kazakhstan.^27-29^ The fake vaccine passports undermine efforts to mitigate COVID-19. Falsification occurs in part because authorities have required COVID-19 vaccinations for certain populations, but vaccine confidence is low. Data on real-word effectiveness of vaccines may help address low vaccine confidence.

Vaccine confidence can also be boosted through increased global recognition of efficacious vaccines. Despite trials showing high vaccine efficacy and studies showing real-world vaccine effectiveness in several countries, only 1 of the 4 vaccines in our study, CoronaVac, is accepted as proof of vaccination in several countries. The other vaccines, 2 of which have been used widely globally, have not completed submission of all requirements for WHO approval.

## Strengths and limitations

This is the first study from Kazakhstan reporting the real-world effectiveness of fours available vaccines based on the retrospective cohort analysis in the largest city of the country using aggregated vaccination data and individual-level breakthrough case national surveillance data. Three of these studied vaccines are not accepted for international travel by most governments worldwide. This is also the first study to estimate vaccine effectiveness for QazVac, an inactivated vaccine developed in Kazakhstan, in real-world conditions. Our results are relevant for planning and scale-up of COVID-19 vaccination efforts in the region.

Our study is subject to at least five important limitations. First, we could not assess vaccine effectiveness against severe disease, hospitalization, and death because individual level data on unvaccinated people who died or were hospitalized were not available for our analysis. Second, we were not able to adjust for potential confounders, such as age or underlying medical conditions, because this data was not available. Our analysis is likely not biased by age because the population proportion ages 60 years and older among vaccinated people (12%) approximates that proportion in the general adult population.^30^ Additionally, the analysis captures 57% of the vaccinated population. Third, when only people with symptoms of COVID-19 test, vaccine effectiveness may be overestimated. However, testing was widely available in Almaty during the study period, with no documented shortages. There were no changes in testing policy over this time, and proof of COVID-19 negative test results were required for participation in several activities. Also, the seven-day moving average of COVID-19 tests tracked the epidemic curve. Fourth, individual data was available only for COVID-19 cases among vaccinated people and cases among unvaccinated people were aggregate. We could not remove potential duplicates from aggregated COVID-19 case data. Fifth, though we attempted to remove fake vaccine passports from the analysis, we cannot ascertain status of all vaccine records and people who were not vaccinated may have been included in the vaccinated population.

## CONCLUSION

We found that 4 vaccines used in Almaty were highly effective against SARS-CoV-2 infection. Our results may help increase vaccine confidence in Kazakhstan and the region. Scale-up of efforts to fully vaccinate all eligible unvaccinated persons and to providers booster doses to persons already fully vaccinated, people >60 years of age and during variant surges, will help reduce SARS-CoV-2 incidence.

## Data Availability

The authors cannot share individual level data due to the national data protection regulations. Deidentified data might be made available for authorized researchers after application to the Ministry of Healthcare of the Republic of Kazakhstan.
The authors confirm that the manuscript is an honest, accurate, and transparent account of the investigation being reported; and that no important aspects of the study have been omitted.
Dissemination to participants and related patient and public communities: The Ministry of Healthcare of the Republic of Kazakhstan and the authors will disseminate the study findings through press releases, a website (https://rk-ncph.kz/ru/), and through social media

## Contributors

DN, RH, MS, GN, AY, DS, AH and AT conceived and designed the study. DN, RH, and GN did the statistical analysis. MS, GN, AY, and AT were responsible for data curation. DN, RH, AH, and DS searched the existing literature. DN, RH, and AH wrote the original draft of the manuscript and all authors reviewed and edited the manuscript for intellectual content. DN and RH prepared data visualization. AY, DS, and AT were responsible for study administration. The corresponding author confirms that all listed authors meet authorship criteria and that no others meeting the criteria have been omitted.

## Ethical approval

The study was deemed non-research public health activity by the MoH of Kazakhstan and the institutional review board of CDC and conducted in accordance with CDC policy (project ID 0900f3eb81e4b259)

## Data sharing

The authors cannot share individual level data due to the national data protection regulations. Deidentified data might be made available for authorized researchers after application to the Ministry of Healthcare of the Republic of Kazakhstan.

The authors confirm that the manuscript is an honest, accurate, and transparent account of the investigation being reported; and that no important aspects of the study have been omitted.

Dissemination to participants and related patient and public communities: The Ministry of Healthcare of the Republic of Kazakhstan and the authors will disseminate the study findings through press releases, a website (https://rk-ncph.kz/ru/), and through social media.

## Provenance and peer review

Not commissioned; externally peer reviewed.

