## Supplementary figures and images for "Effectiveness of Four Vaccines in Preventing SARS-CoV-2 Infection in Kazakhstan"

### Supplementary figure

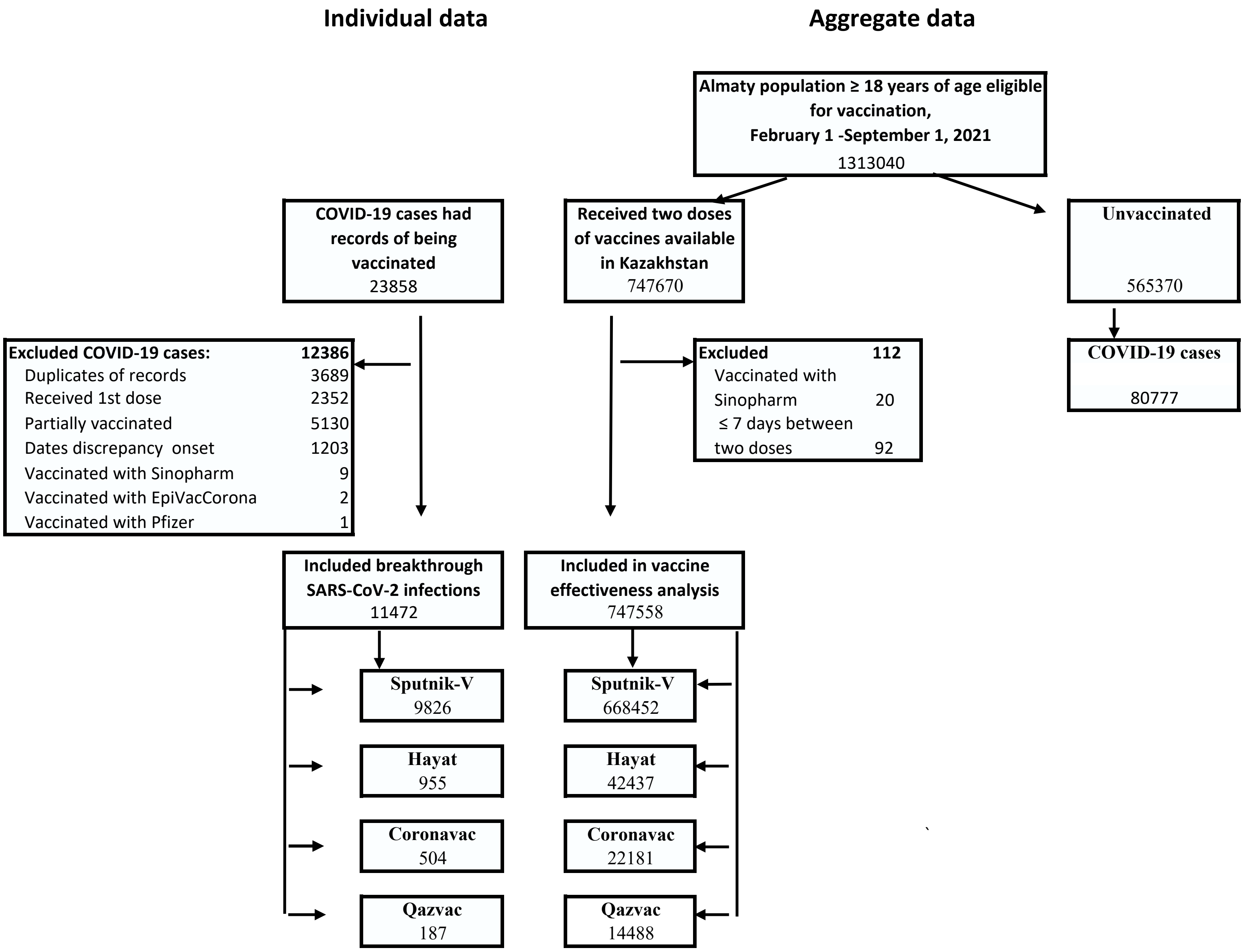
